# Sources of variability in methods for processing, storing, and concentrating SARS-CoV-2 in influent from urban wastewater treatment plants

**DOI:** 10.1101/2021.06.16.21259063

**Authors:** Joshua A. Steele, Amity G. Zimmer-Faust, John F. Griffith, Stephen B. Weisberg

## Abstract

The rapid emergence of wastewater based surveillance has led to a wide array of SARS-CoV-2 RNA quantification methodologies being employed. Here we compare methods to store samples, inactivate viruses, capture/concentrate viruses, and extract/measure viral RNA from primary influent into wastewater facilities. We found that heat inactivation of the viruses led to a 1-3 log_10_ decrease compared to chemical inactivation. Freezing influent prior to concentration caused a 1-4 log_10_ decrease compared to processing fresh samples, but viral capture by membrane adsorption prior to freezing was robust to freeze-thaw variability. Concentration vs. direct extraction, and PCR platform also affected outcome, but by a smaller amount. The choice of nucleocapsid gene target had nearly no effect. Pepper mild-mottle virus was much less sensitive to these methodological differences than was SARS-CoV-2, which challenges its use as a population-level control among studies using different methods. Better characterizing the variability associated with different methodologies, in particular the impact of methods on sensitivity, will aid decision makers in following the effects of vaccination campaigns, early detection of future outbreaks, and potentially monitoring the appearance of SARS-CoV-2 variants in the population.

## 1. Introduction

Wastewater based surveillance (WBS) of SARS-CoV-2 RNA is gaining traction because of its many advantages over individual testing, particularly the cost-effectiveness of a relatively unbiased pooled sample and the ability to detect virus shed from infected asymptomatic or pre-symptomatic individuals (Bivins et al. 2020, Hart & Halden 2020, Kitajima et al. 2020, Ahmed et al. 2021). WBS also yields information several days and almost two weeks faster than it takes to collate individual testing and hospitalization records, respectively (Nemudryi et al. 2020). As a result, SARS-CoV-2 RNA is being measured in the sewage influent stream throughout the world (Medema et al. 2020, Ahmed et al. 2020a, Gerrity et al. 2021, Graham et al. 2021, Gonzalez et al. 2020), from more than 2000 sites in 50 countries (Naughton et al. 2021). The application of WBS to SARS-CoV-2 builds upon the success of WBS for monitoring other viral pathogens, including poliovirus, hepatitis A & E, rotavirus, adenovirus, and norovirus (Katayama et al. 2008, Ashgar et al. 2014, Alleman et al. 2021, McCall et al. 2020).

The rapid emergence of WBS for SARS-CoV-2, though, has led to a wide array of quantification methodologies being employed (Farkas et al. 2020). With little known about the SARS-CoV-2 virus, laboratories had to make decisions about how to inactivate it in samples prior to processing so as to meet safety guidelines. Decisions about extraction and concentration techniques, and whether to use qPCR vs. ddPCR, were largely made based on existing practices with other pathogens within each laboratory, as there was insufficient lead time to measure the effects of such techniques. Target gene selection was often made on availability of primer sets that were initially in scarce supply. Laboratories also had to determine quickly whether to process samples fresh or hold them frozen while they investigated methodological processing details.

Several studies have looked at the effects of those decisions, the largest of which included 32 laboratories processing two sets of common samples (Pecson et al. 2021). That study found up to 7 orders of magnitude difference in SARS-CoV-2 concentrations across laboratories when the same samples were processed using different methods, which was considerably larger than within method variability. However, disentangling confounded method effects from such large studies is challenging. There have been a few controlled experiments looking at specific methodological choices, such as concentration techniques (Ahmed et al. 2020b, Torii et al. 2020, La Rosa et al. 2021, LaTurner et al. 2021, Whitney et al. 2021) and processing platform (Feng et al. 2021, Cieselski et al 2021, Ahmed et al. 2021). Here we expand upon those efforts by conducting controlled experiments to examine a range of sample processing choices (Fig. 1).

**Figure 1.**
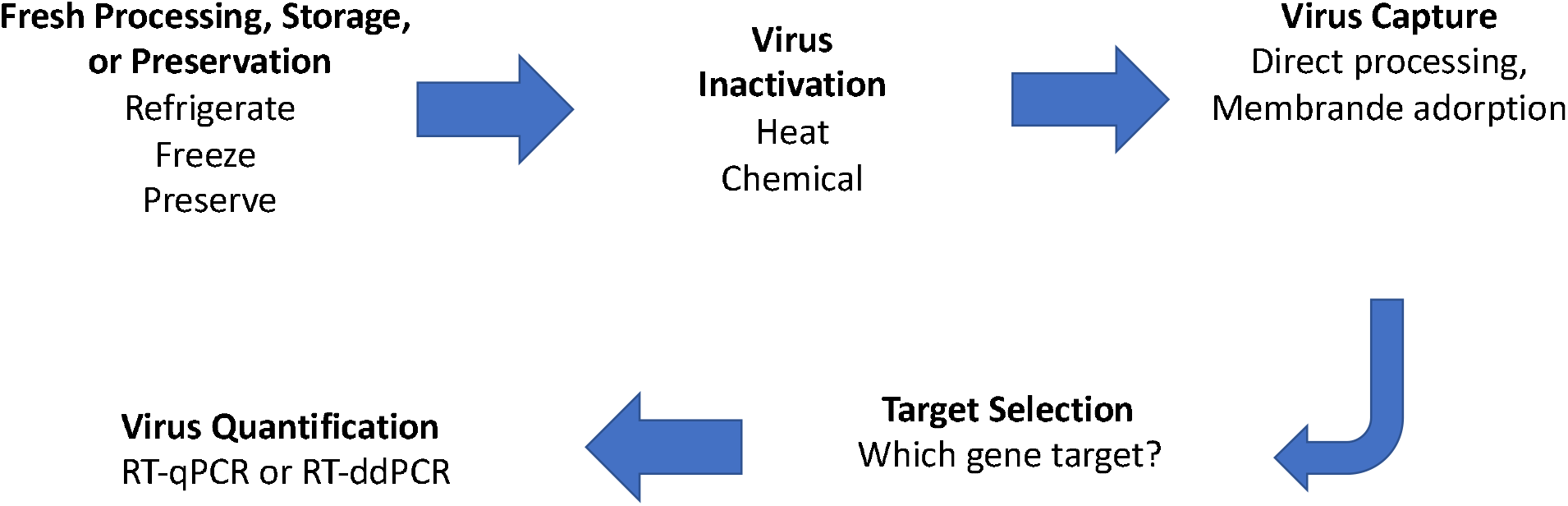
Virus in wastewater sample processing steps that were evaluated in this study

## 2. Materials and Methods

### 2.1 Experimental Design

Controlled experiments were performed in two parts (Figure 1). The first set of comparisons tested the effects of freezing and sample storage, heat inactivation, and direct extraction vs concentration on a membrane filter using triplicate influent samples from multiple Southern California WWTPs. For this set of experiments each wastewater sample was aliquoted and processed in parallel three different ways described below (section 2.2). Briefly, 1) the sample was concentrated on a mixed cellulose ester membrane and extracted using bead beating lysis followed by magnetic bead capture; 2) the sample was preserved in lysis buffer and then extracted using bead beating lysis followed by a total nucleic acid magnetic bead capture, and 3) the sample was preserved in DNA/RNA Shield and then extracted using a bead beating lysis followed by silica spin column RNA extraction.

The second set of comparisons tested the SARS-CoV-2 concentrations measured by two widely used nucleocapsid gene assays (N1: 2019 nCOV N1, N2: 2019 nCOV N2, Table S2) using 296 influent samples from three southern California WWTPs. Thirty of these samples from one WWTP were also used to compare RT-QPCR and RT-droplet digital PCR platform for the SARS-CoV-2 N1 gene. For these comparisons the wastewater sample was concentrated on a mixed cellulose ester membrane then extracted using bead beating lysis followed by magnetic bead capture.

All comparisons took place on homogenized wastewater samples from five WWTP in Southern California, USA collected and transported to the laboratory at 4°C. Individual WWTP used for each comparison are described in the Supplemental Material (Table S1).

#### 2.1.1 Sample Storage and Preservation

Wastewater samples were processed and preserved four different ways. First, samples were immediately processed, extracted, and analyzed via RT-ddPCR on the same day (treatment designation: fresh). Secondly, samples were immediately amended with HCL and MgCl_2_ and concentrated on a membrane or amended with lysis buffer (DE:BM)) or DNA/RNA shield (DE:Z) and stored at −80°C for at least 24 hours before thawing, extraction and analysis via RT-ddPCR (fresh processed). Third, raw influent was frozen at −80°C for at least one week, then thawed at 4°C and processed following the same protocol as the Fresh samples (frozen). Lastly, raw influent was frozen at −80°C for at least one week and processed following the same protocol as the Fresh Processed samples (frozen X2).

#### 2.1.2 Inactivation of Viruses by Heat (Pasteurization)

Homogenized wastewater samples were processed two different ways: placed in a water bath at 70°C for 40 minutes or kept at 4°C. For each treatment, a set of samples was concentrated by membrane adsorption and extracted using the bioMerieux magnetic bead extraction kit (HA), or extracted directly from wastewater using the bioMerieux magnetic bead extraction kit (DE:BM) or the Zymo Microbiomics kit (DE:Z).

#### 2.1.3 Virus Capture and RNA Extraction

Wastewater samples were collected during two different time periods, to evaluate differences between virus capture methods during different levels of background SARS-CoV-2 in the human population. Samples were collected in August 2020, during a period of lower SARS-CoV-2 levels in wastewater, and December 2020, during a period of higher SARS-CoV-2 levels in wastewater.

#### 2.1.4 SARS-CoV-2 Gene Target Selection and comparison of gene quantification using RT-QPCR and RT-ddPCR

The concentrations of two widely used nucleocapsid gene assays (CDC-N1, CDC-N2) in influent from 296 samples across three southern California WWTPs were compared. These samples were processed with the membrane adsorption method (HA) with acidification and MgCl_2_ addition (Section 2.7). A subset (n = 30) of these samples from one wastewater treatment plant was analysed for SARS-CoV-2 using both RT-QPCR and RT-digital PCR for comparison.

### 2.2 Sample Processing

#### 2.2.1 Virus Adsorption to Mixed Cellulose Ester Filters and Magnetic Bead Extraction (HA)

Prior to filtration, bovine coronavirus that was obtained as Bovilis® bovine coronavirus vaccine (Merck & Co Inc, Kenilworth, NJ) was added as a sample processing control for assessing SARS-CoV-2 viral RNA recovery. To collect viruses, wastewater samples were amended to a final concentration of 25 mM MgCl_2_ and pH of <3.5 through addition of 20% HCl and concentrated on a mixed cellulose ester membrane (type HA: Millipore, Bedford, MA). Samples were filtered in replicate (n=6) for each site and sampling day with 20 mL wastewater concentrated per filter.

Nucleic acid was extracted the same day using bead beating lysis. HA filters were transferred to pre-loaded 2 mL ZR BashingBead Lysis tubes (Zymo, Irvine, CA, USA) along with the 600 µl NucliSENS lysis buffer. Bead beating was carried out on the Biospec beadbeater (Biospec Products, Bartlesville, OK) for 1 minute. After bead beating, total nucleic acids were extracted using the bioMerieux NucliSENS extraction kit and magnetic bead capture (bioMerieux NulciSENS) according to the manufacturer’s instructions.

#### 2.2.2 Direct Virus Nucleic Acid Extraction via Magnetic Bead Extraction

Total nucleic acid was extracted by transferring 750 µl of homogenized raw influent into pre-loaded 2 ml ZR BashingBead Lysis tubes (Zymo Research, Irvine, CA, USA) along with 750 µl NucliSENS lysis buffer and bead beating was carried out on the Biospec beadbeater (Biospec Products, Bartlesville, OK) for 1 minute. After bead beating, total nucleic acids were extracted using the bioMerieux NucliSENS extraction kit and magnetic bead capture (bioMerieux NulciSENS) according to the manufacturer’s instructions.

#### 2.2.3 Direct Virus Nucleic Acid Extraction via Silica Column

Total nucleic acid was extracted by transferring 250 µl of homogenized raw influent into pre-loaded 2 mL ZR BashingBead Lysis tubes (Zymo Research, Irvine, CA, USA) along with 1.2 ml of DNA/RNA shield (Zymo Research, Irvine, CA). Bead beading was carried out on the Biospec beadbeater (Biospec Products, Bartlesville, OK) for 1 minute. After bead beating, total nucleic acids were extracted using the Zymo Microbiomics RNA Extraction kit (Zymo Research, Irvine, CA) according to the manufacturer’s instructions.

### 2.3 Virus Quantification via RT-ddPCR

Extracted RNA was analyzed using reverse transcriptase droplet digital PCR (RT-ddPCR) for the N1 and N2 regions of the SARS-CoV-2 N gene, Pepper Mild Mottle Virus (PMMoV), and Bovine Coronavirus (BCoV) genes using the One-Step RT-ddPCR Advanced Kit for Probes (Bio-Rad, Hercules, CA). Primer and probe sequences for SARS-CoV-2 N1 and N2 were those designed by the United States Center for Disease Control (CDC; Lu et al. 2020), PMMoV (Kitajima et al. 2018, Gonzalez et al. 2020), and BCoV (Decaro et al. 2008) are described in table S2. Each primer was added at a final concentration of 0.9 µM and probes were added at a final concentration of .25 µM. 5 µl of RNA extract was added to each reaction for a final volume of 20 µl. Plates were placed into a Bio-Rad C1000 Touch thermocycler (Bio-Rad, Hercules, CA) and underwent reverse transcription at 50°C for 1 hour. Enzyme activation and initial denaturation were performed at 95°C for 10 minutes, then 40 cycles of denaturation at 95°C for 30 seconds, annealing/extension at 58°C for 1 minute. Enzyme deactivation was performed at 98°C for 10 minutes followed by a 12°C hold for 20 minutes before being placed in the QX200 (Bio-Rad, Hercules, CA) for droplet reading. For all assays, a minimum of two reactions and a total of ≥20,000 droplets were generated per sample and at least five no template control (NTC) reactions and two positive control reactions were run per 96-well plate as well as extraction-specific NTCs. To consider a sample positive, quantifiable, and included in further analysis, each sample was required to have a minimum of three positive droplets which served as the threshold for the limit of quantification (Cao et al. 2015, Steele et al. 2018). The limit of quantification was converted using the following equation:

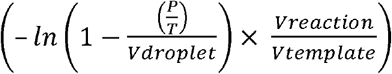

Where *P* is the number of positive droplets, *T* is the total accepted droplets, *V*_*droplet*_ is the average volume per droplet expressed in microliters (i.e. ∼0.0009 µl which is equal to ∼0.9 nanoliters), *V*_*reaction*_ is the volume of the digital PCR reaction, and *V*_*template*_ is the volume of sample extract (PCR template) added to the reaction. This per µl template reaction can then be converted to copies per ml by multiplying by the volume of sample extract divided by the volume of sample filtered (Steele et al. 2018). RNA recovery was also assessed using the BCoV exogenous control. Samples where recovery fell below 3% were excluded from further analyses.

### 2.4 Virus Quantification via RT-qPCR

One-step reverse transcription quantitative PCR (RT-qPCR) was used to quantify the N1 and N2 regions of the SARS-CoV-2 N gene. Bio-Rad iTaq Universal Probes One-Step Kit was used according to the manufacturer’s instructions (Bio-Rad, Hercules CA). Each primer was added at a final concentration of 0.9 µM and probes were added at a final concentration of 0.25 µM. 5 µl of RNA extract was added to each reaction for a final volume of 20 µl. The IDT 2019-nCoV_N_Positive Control plasmid (IDT, San Diego, CA) was used to make a standard curve. The plasmid was linearized using Xmn1 restriction enzyme in 1X rCutSmart™ Buffer (New England BioLabs, Ipswich, MA) at 37°C for 1 hour followed by denaturation at 65°C for 20 minutes. A 6-point standard curve was created by diluting the linearized plasmid covering a range from 10^6^-10^1^ copies per reaction. Plates were placed into a CFX 96 Touch thermocycler (Bio-Rad, Hercules, CA) and underwent reverse transcription at 50°C for 10 minutes. Enzyme activation and initial denaturation were performed at 95°C for 3 minutes, then 40 cycles of denaturation at 95°C for 15 seconds, annealing/extension at 58°C for 30 seconds. To consider a sample positive and included in further analysis, both reactions The limit of quantification for the wastewater was calculated to be 1000 copies per 100ml sample based on the lowest Cq value obtained on the standard curve: 34.5. The standard curve regression equation was 3.205x+37.26 (r^2^=0.997) with an efficiency of 105.1%. All NTCs did not amplify.

### 2.5 Data Analysis and Statistics

Statistical analyses throughout this report were conducted in R (R Core Team 2020), utilizing log_10_-transformed concentrations. Analysis of Variance (ANOVA) tests were conducted to assess for significant differences in concentration among the different experimental treatments. Separate ANOVA tests were completed for each target measured (SARS-CoV-2 N2, SARS-CoV-2 N2, PMMoV). When a significant difference was found, the multcomp package in R (Hothorn et al. 2008) was used to run post hoc Tukey comparisons for individual pairwise comparisons between the different treatment levels.

## 3. Results

### 3.1 Freezing of influent and preserved or concentrated samples

Freezing Influent samples at −80°C (frozen treatment) and processing through direct extraction resulted in 1-5 log_10_ lower recoveries compared to fresh influent kept for less than one day at 4°C (fresh treatment), while influent samples processed using membrane adsorption showed little effect of freezing at ultra-low temperatures. Freezing the influent at −80°C resulted in a significant reduction of the SARS-CoV-2 N1 and N2 concentrations by direct extraction for both the Zymo (DE:Z) and the bioMerieux kits (DE:BM) of approximately 2-4 Log_10_ and more than 4 Log_10_, respectively, to samples held at 4°C and processed the same day. For the bioMerieux kit, SARS-CoV-2 N1 (F=22.13, all pairwise p-values <0.001) and SARS-CoV-2 N2 (F=91.63, all pairwise p-values<0.001) concentrations were significantly lower for all frozen samples. For the Zymo kit, SARS-CoV-2 N1 (F=4.384, p<0.05) and SARS-CoV-2 N2 (F=4.562, all pairwise p-values <0.05) concentrations were significantly lower for the frozen X2 samples only. In contrast, freezing the influent resulted in no significant change in both SARS-CoV-2 N1 (F=1.418, all pairwise p-values >0.05) and SARS-CoV-2 N2 concentrations (F=2.009, all pairwise p-values >0.05) for samples concentrated by membrane adsorption (Fig. 2).

**Figure 2.**
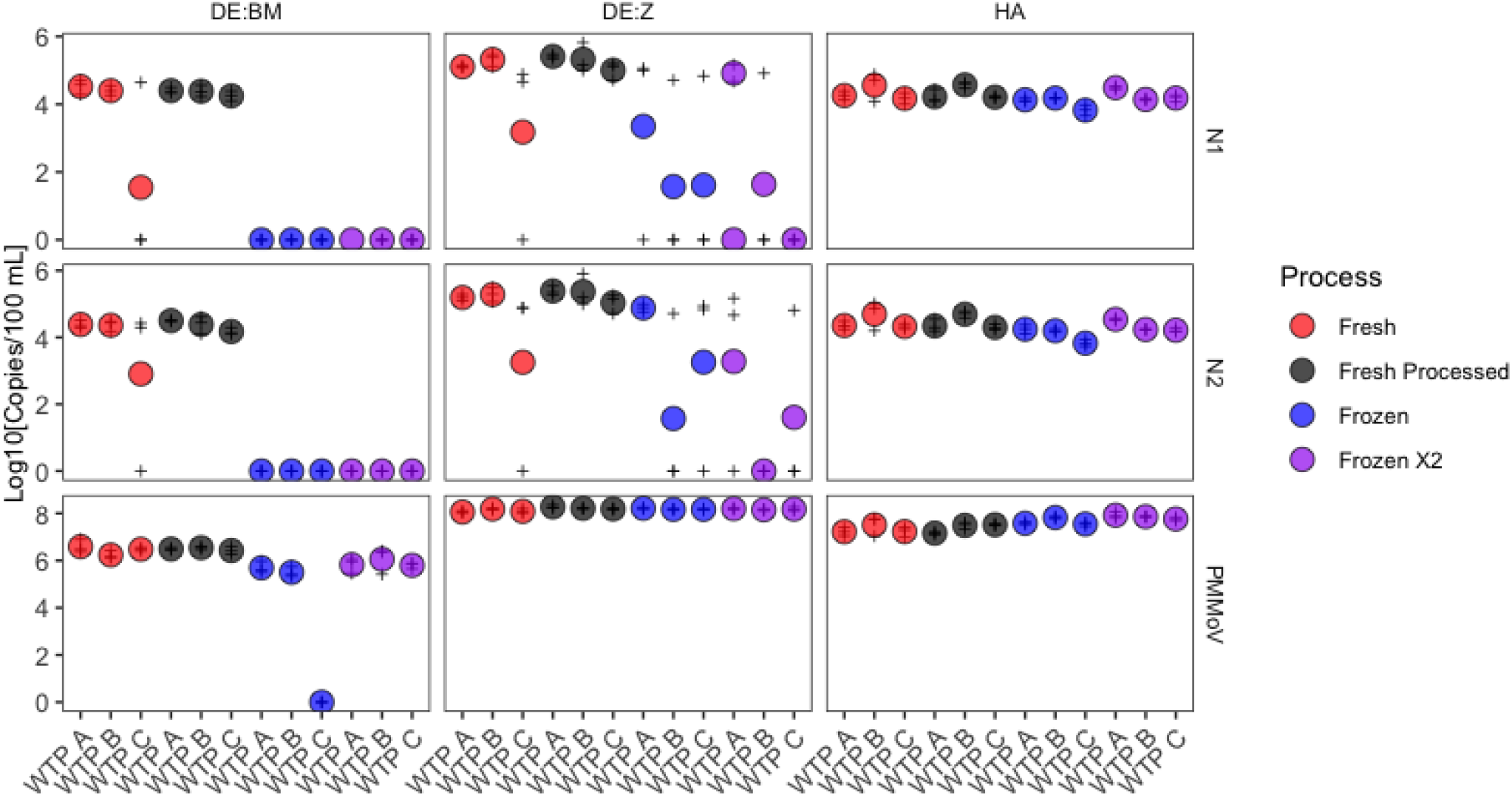
Comparison of frozen storage methods using samples from 3 WWTP by direct extraction methods (DE:BM & DE:Z) and membrane adsorption (HA) prior to extraction. Concentrations are reported in log_10_ copies per 100ml for N1 (top row), N2 (bottom row), and PMoMV (bottom row). Circles represent average concentration for the three WTPs and faint crosses represent results from the individual plants. Fresh represents samples with no freeze-thaw, Fresh Processed represents fresh influent samples chemically preserved or concentrated on a membrane prior to being frozen, Frozen represents influent that has gone through one freeze-thaw cycle prior to preservation or concentration, Frozen X2 is influent that has gone through one freeze thaw, then is preserved or concentrated on a membrane, then goes through a second freeze thaw prior to extraction.

Samples which were preserved, then frozen (fresh processed and frozen X2) and then put through a direct extraction resulted in similar concentrations as the samples which did not undergo a second freeze-thaw. Influent held at 4°C, preserved, then frozen (fresh processed) for both the DE:BM and DE:Z direct extraction samples, had concentrations that were nearly the same as those where the preserved sample was stored at 4°C: 4-5 Log_10_ SARS-CoV-2 N1 and N2 copies per 100ml. Samples which were frozen, thawed, preserved, then frozen again (frozen X2) for influent stored at 4°C and below detection for frozen influent. SARS-CoV-2 N1 and N2 concentrations varied less than 0.5 Log_10_ with the addition of a freeze-thaw step for influent adsorbed onto a membrane filter.

All three methods yielded quantifiable PMMoV concentrations when the influent was stored at 4°C and no impact was observed with freezing the preserved of concentrated samples, with the exception of one WTP plant (Fig. 2). Concentrations of PMMV varied less than 0.5 Log_10_ at 4°C or after a freeze thaw.

### 3.2 Heat Inactivation of Viruses

Heat inactivation at 70°C resulted in a significant reduction, between 1-3 Log_10_, in SARS-CoV-2 N1 (F=107.1, P<0.001) and N2 (F=3347, P <0.001) concentrations for membrane concentrated samples (Figure 3A). The SARS-CoV-2 N1 and N2 concentrations were below the limit of detection for all but one untreated or heat inactivated direct extraction. Therefore, BCoV spike-in recovery concentrations were compared instead for direct extraction using both the Zymo (DE:Z)and bioMerieux kits (DE:BM) (Figure 3B).

**Figure 3.**
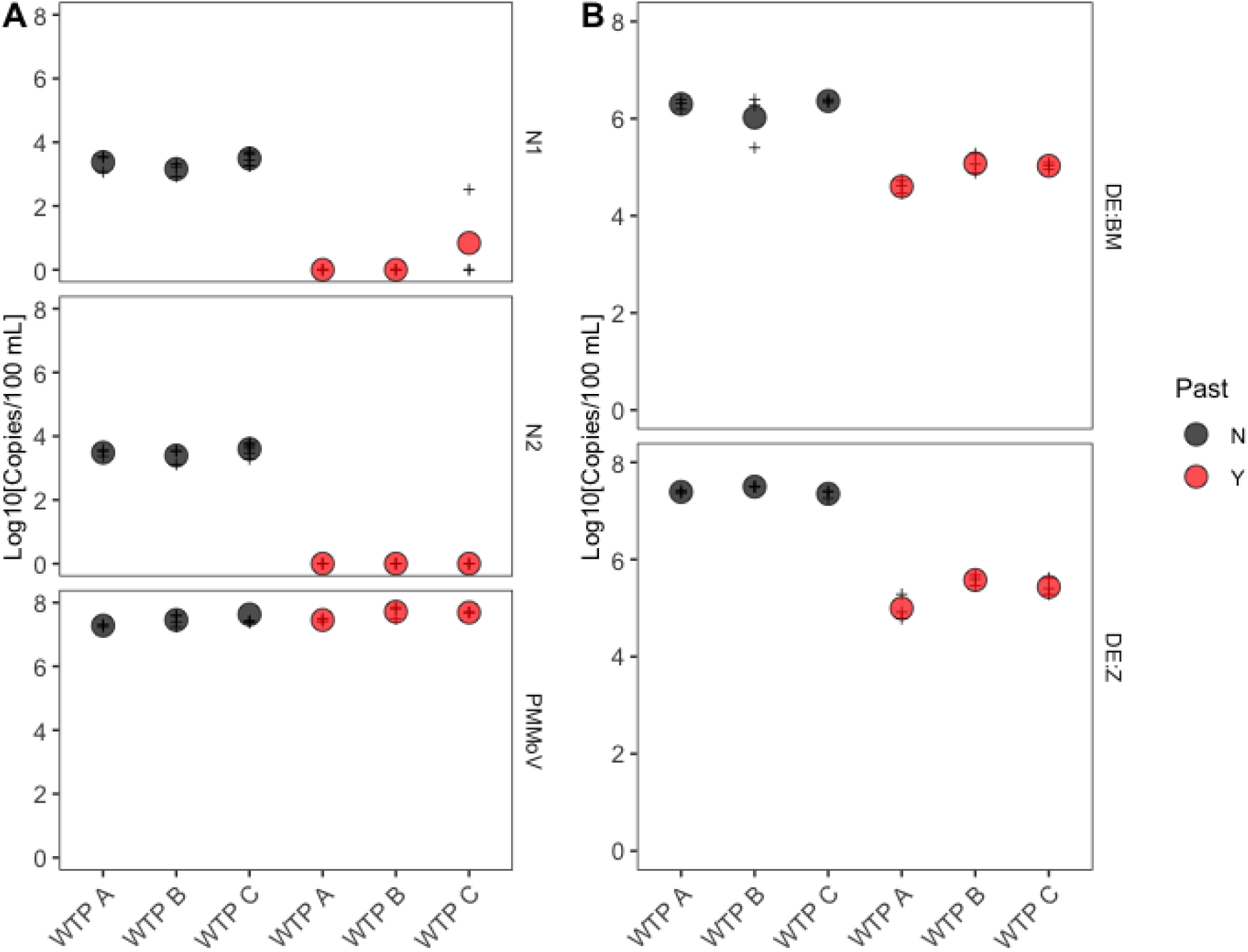
Concentrations measured with and without pasteurization. Circles represent average concentration for the three WTPs and faint crosses represent results from the individual plants. Black circles indicate samples not pasteurized; red circles indicate pasteurized samples; (A) SARS-CoV-2 N1 (top row), SARS-CoV-2 N2 (middle row), and PMMV (bottom row) levels for samples processed by membrane adsorption (HA) B) BCoV levels by direct extraction methods (DE:BM & DE:Z).

Heat inactivation at 70°C resulted in a significant reduction, approximately 2 Log_10_, in BCoV levels for samples processed via direct extraction using the Zymo kit (F=132.3, P<0.001). For samples processed by direct extraction using the bioMerieux, BCoV levels were reduced by approximately 1-2 Log_10_ (F=52.36, P<0.01, Figure 3). In contrast to SARS-CoV-2 and BCoV, PMMoV concentrations did not significantly decrease following heat inactivation at 70°C for 40 minutes (F=1.571, P>0.05, Figure 3).

### 3.3 Direct Extraction vs Membrane Adsorption

Membrane adsorption enabled quantification of viral RNA over a wider range of concentrations compared to either direct extraction method. Direct extraction using either the bioMerieux (DE:BM) or Zymo kit (DE:Z) were unable to reliably recover SARS-CoV-2 RNA when the concentrations were low: 10^3^ − 10^4^ copies per 100ml (Figure 4A). Only the wastewater samples from WTP C had a quantifiable amount in two of the triplicate samples. The other wastewater samples were all below detection. In contrast, concentration by membrane adsorption (HA) resulted in measurable, and significantly higher, concentrations for both N1 (F=10.2, all pairwise P-values <0.05) and N2 (F=15.06, all pairwise P-values <0.01) when SARS-CoV-2 concentrations were 10^3^ − 10^4^ copies/100 mL in wastewater.

**Figure 4.**
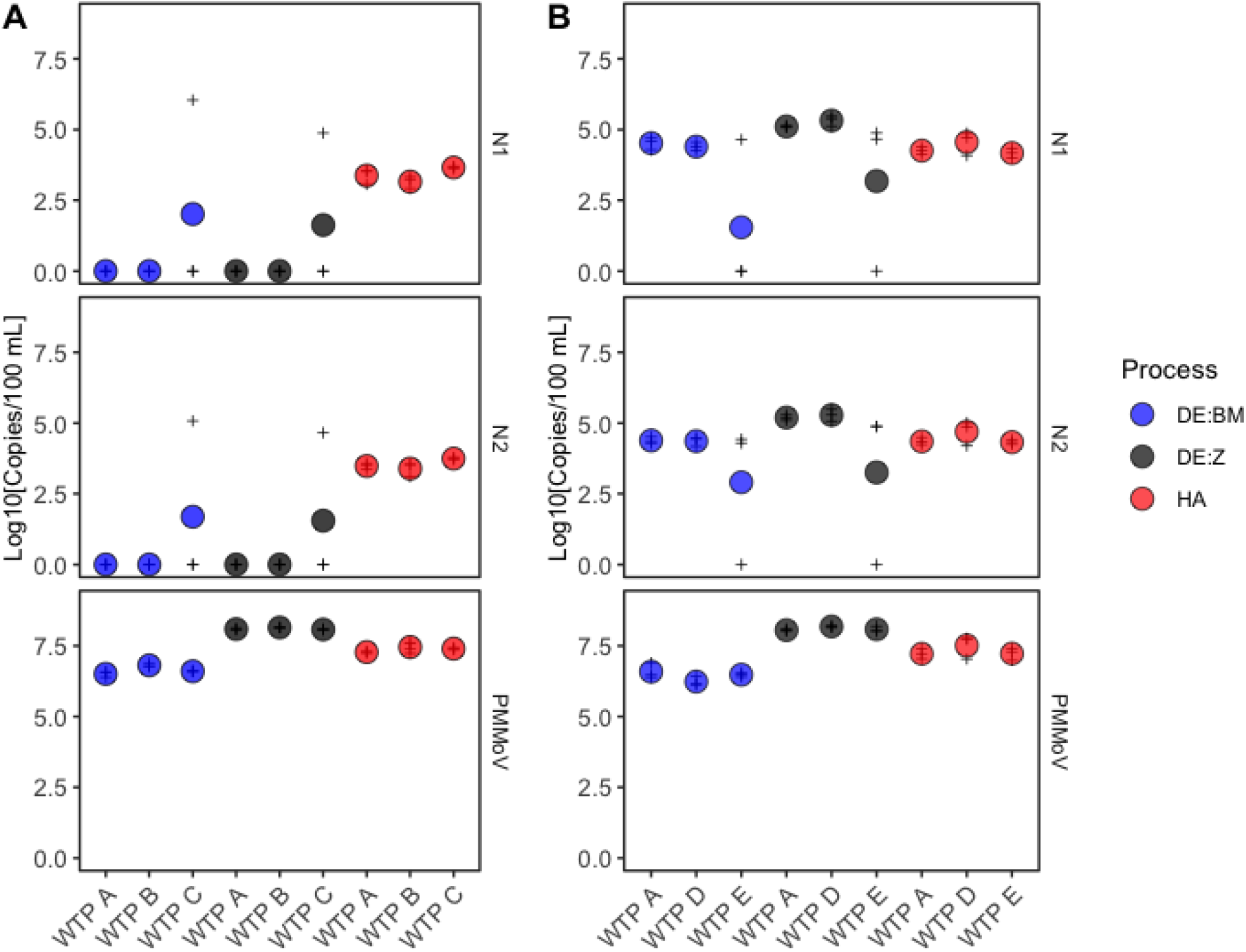
Differences between concentration methodology. Color of the circles indicate method used. HA: membrane adsorption; DE:BM direct extraction with the bioMeriuex kit; DE:Z: direct extraction with the Zymo kit. (A) N1 and N2 levels measured August 2020, during lower levels of SARS-CoV-2 in wastewater. B) N1 and N2 levels measured December 2020, during higher levels of SARS-CoV-2 in wastewater. Circles represent average concentration for the three WTPs and faint crosses represent results from the individual plants.

At SARS-CoV-2 concentrations of 10^4^-10^5^ copies per 100 mL, direct extraction by both the bioMerieux (DE:BM) and Zymo kits and membrane adsorption (HA) provided similar concentrations of SARS-CoV-2

N1 (F=0.65, all pairwise P-values >0.05) and N2 (F=0.59, all pairwise P-values >0.05) copies and overall direct extraction was able to more reliably quantify SARS-CoV-2 N1 and N2 copies, than at the lower concentrations measured (Figure 4B). However, membrane adsorption method (HA) was still the only method for which all replicates produced quantifiable results.

All methods were able to produce quantifiable results for PMMOV (Figure 4). Direct extraction using the Zymo kit (DE:Z) yielded the highest PMMOV copies followed by membrane adsorption (HA), and direct extraction using the bioMerieux kit (DE:BM).

### 3.4 Comparison of N1 vs N2 gene targets

The N1 and N2 assays produced similar SARS-CoV-2 concentration measurements in wastewater from 3 Southern California treatment plants (Fig 5). The measurements were highly correlated (R^2^ =0.95, p<0.001). The slope of the regression line was 1.28, reflecting the generally higher concentrations observed by the N2 assay.

**Figure 5.**
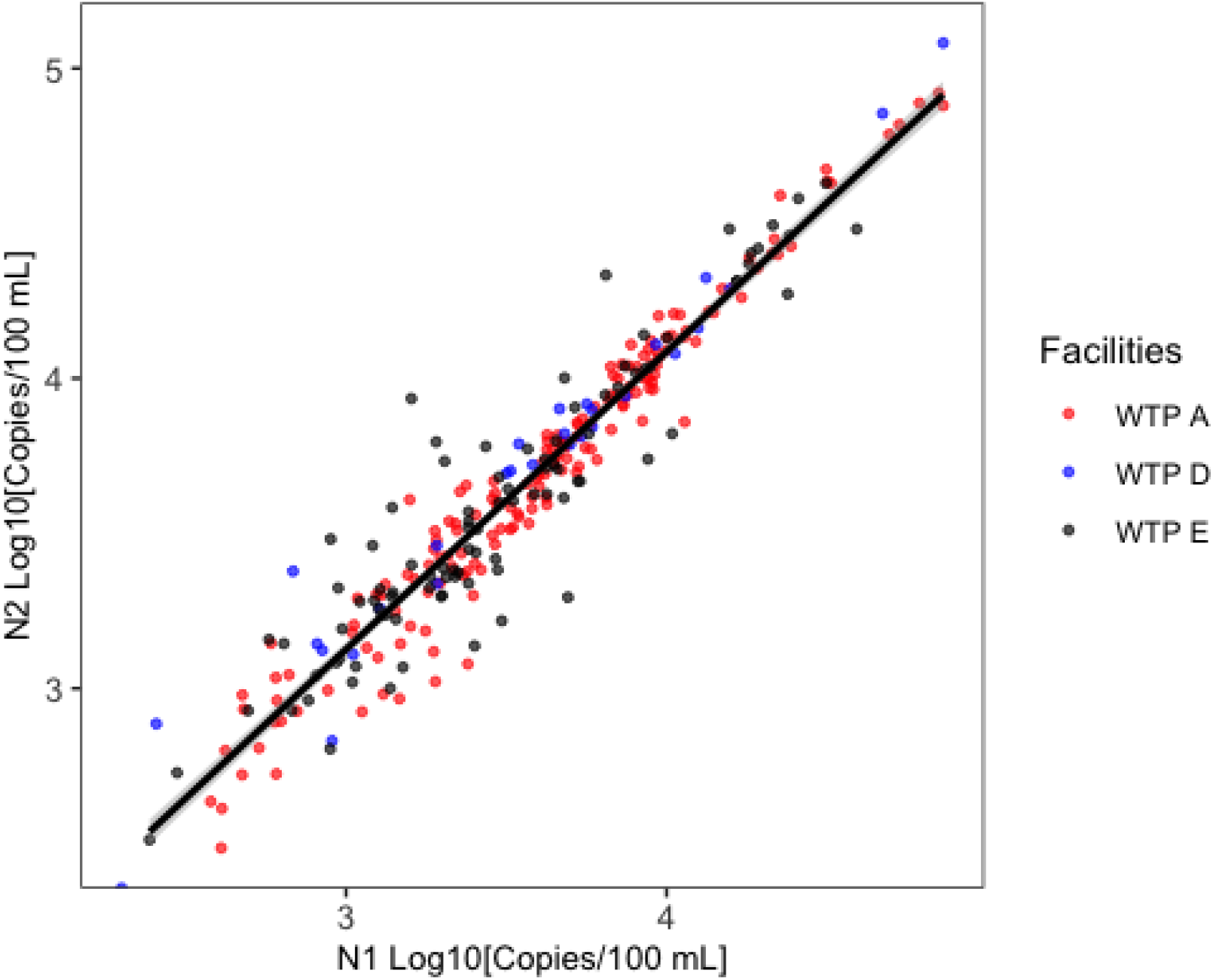
Log10 SARS-CoV-2 in wastewater from three sewage treatment plants measured using the SARS-CoV N1 and N2 assays.

### 3.5 Comparison of RT-qPCR to Digital PCR

SARS-CoV-2 N1 gene concentrations measured by digital droplet RT-PCR (RT-ddPCR) and RT-qPCR (Fig 6) were highly correlated (R^2^ =0.80, p<0.001), though there was a slight tendency to higher measurements in RT-qPCR (regression slope of 1.68). The correlation declined in samples with lower SARS-CoV-2 concentration. In addition, RT-ddPCR produced detectable, quantifiable concentrations in all 30 samples, but RT-qPCR had 3 samples where the gene was not detected and 3 additional samples that were below the limit of quantification.

**Figure 6.**
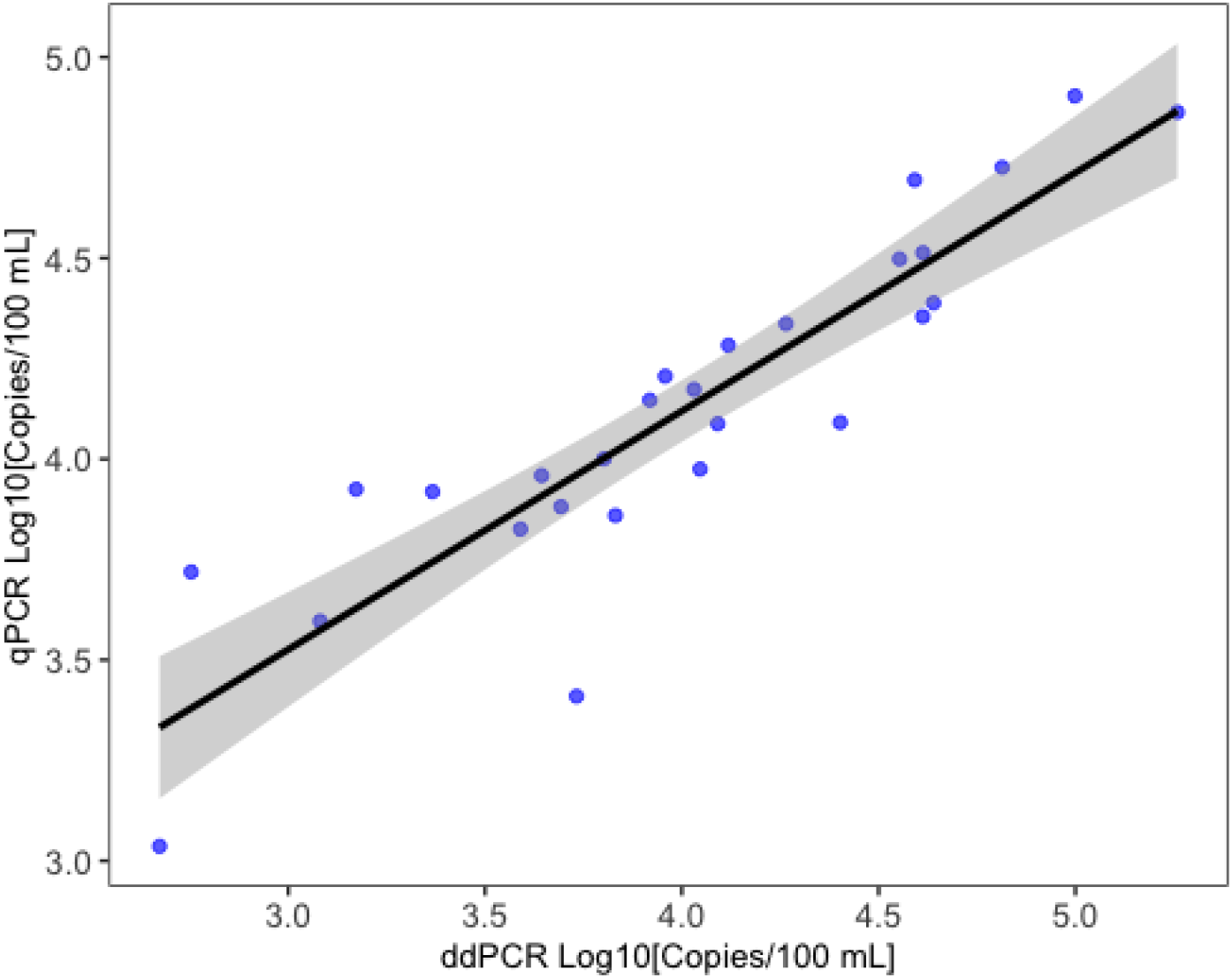
Log10 SARS-CoV-2 N1 in wastewater measured using qPCR (y-axis) and digital PCR (x-axis).

## 4 Discussion

Our finding that freezing/thawing of whole influent samples can substantially reduce measured concentrations of SARS-CoV-2 is largely, but not entirely, consistent with other studies that have examined this effect. Wiedhaas et al. (2020) found a 92% reduction in SARS-CoV-2 following freezing influent at ultra-low temperatures for one week prior to processing. Using coronavirus OC43, McMinn et al. (2021) found up to 0.5 log_10_ reduction after freezing virus solutions for one week at ultra-low temperatures. This result differs from Hokajärvi et al. (2021), who found little reduction from freezing/thawing influent sample using ultra-low freezer temperatures.

The effect of freezing appears to be easily mitigated in several ways. We found that removing the water matrix and capturing virus on charged filters prior to freezing mitigated this effect. Adding a chemical preservative prior to freezing (e.g., salt, PEG, or an elution solution) has also been reported to reduce viral decay (Whitney et al. 2021, Pecson et al. 2021, McMinn et al. 2021). The observed benefits from adding additional solute particles to the water matrix to mitigate viral decay are consistent with results reported when another solute rich matrix, primary sludge, has been frozen (Graham et al. 2021, Simpson et al. 2021).

Our finding that measurement using qPCR and ddPCR are highly correlated is consistent with other studies in nasopharyngeal swabs (Falzone et al. 2020, Liu et al. 2020), plasma (Tedim et al. 2021), wastewater from aircraft and cruise ships (Ahmed et al. 2020), and raw influent (Cieselski et al. 2021). However, we observed that the correlation became weaker at the lower end of the concentration range we tested, spreading out at lower concentrations in a “broom-shaped” pattern consistent with studies comparing measurements of bacterial targets (Cao et al. 2015). This difference in sensitivity between measurement methods is consistent with ddPCR having up to a 200X lower limit of detection than does qPCR (Cieselski et al. 2021, Ahmed et al. 2021). Furthermore, the digital PCR platform allows for even lower limits of detection by increasing the number of reactions and the number of droplets measured, although at an increase in reaction materials and cost (Huggett et al. 2014). Sensitivity of RT-qPCR could be improved by increasing the volume of RNA extract, but that also increases the amount of inhibitory compounds which come along in wastewater and have been shown to reduce sensitivity when concentrations are low (D’Aoust et al., 2021; Gerrity et al. 2021, Gonzalez et al., 2020; Graham et al., 2021). While not completely immune, digital PCR has been shown to be more robust to inhibition (Cao et al. 2015).

Our finding that gene target had little effect reinforces several previous reports (e.g. Gerrity et al. 2021, Gonzalez et al 2020, Feng et al. 2021). Even the groups that reported statistically different results among targets (e.g., Pecson et al. 2021) found the difference was less than 10%, much smaller than the other methodological differences we studied. However, our results differ from those of Randazzo et al. (2020) in that we observed that the results from the N1 and N2 gene targets were highly correlated (0.95 vs. 0.5). The reason for this difference is likely due to Randazzo measuring low concentration samples using Rt-qPCR; our use of RT-ddPCR helps ameliorate the increasing variability seen in qPCR results as target concentrations near the theoretical limit of quantification.

Previous studies have been less consistent regarding the effect of heat inactivation (pasteurization). In particular, Pecson et al. (2021) found that pasteurization even caused a slight increase in concentration for some methods (e.g. PEG precipitation), after correction for controls. We found that it caused a 10-1000X decrease, but that may be specific to performing direct extraction with no concentration step. Other authors have found this is of less concern when concentrating samples using PEG precipitation (Pecson et al. 2021) or when NaCl is added prior to performing RNA extraction using silica milk (Whitney et al. 2021). These concentration techniques both use chemicals which will trap viruses, proteins, and nucleic acids (McSharry & Benzinger 1970, Polson 1970, Yamamoto et al. 1970, Lewis and Metcalf 1998) and are using centrifugation to pellet the material which may reduce the loss from virus capsid disruption during pasteurization. In contrast, the membrane adsorption (HA) method uses charge to capture virus particles on an electronegative membrane in the presence of cations and may allow free viral RNA to pass through (Katayama et al. 2002).

One of our more interesting findings was the tradeoff associated with concentration vs. direct extraction. We found that direct extraction had better recovery compared to concentration on a membrane, but a poorer limit of detection. This is consistent with other studies that have looked at concentration vs direct extraction finding a tradeoff in recovery (Ahmed et al. 2020b, Rusiñol et al. 2020, Pecson et al. 2021). This is likely due to the difference in volume that could be processed by the direct extraction (< 0.5ml) compared to concentration (20ml). While Pecson et al (2021) found some of the highest recoveries from small volume direct extraction, Ahmed et al. (2020) identified HA filtration with cation addition as a good tradeoff of recovery and concentration and Gonzalez et al. (2020) successfully applied this technique to monitor multiple WWTPs. As such, small volume direct extraction without concentration may not be best option with lower SARS-CoV-2 concentrations.

Our findings appear to have a number of implications for application of WBS. First and foremost, our findings reinforce Pecson et al.’s (2021) suggestion that the same method should be retained when assessing SARS-CoV-2 concentrations over time at a given location. The several orders of magnitude range of different methodological responses we observed were as large as the entire range of values observed at the facilities from which we collected the samples from in this study (Figs 5, 6).

The second is that methodological differences challenge the ability to make geographic comparisons across facilities that use different methods. Cross-system comparisons are challenging even when using the same methods, as differences in sources among sewersheds (e.g., residential vs. industrial) alters the relationship between RNA and population infectivity, as does transit-time induced differences in decay. A number of studies have attempted to correct for these sewershed difference by normalizing to Pepper Mild Mottle Virus (PMMoV). Unfortunately, our finding that PMMoV is differentially affected by method permutations compared to SARS-CoV-2 makes that correction potentially problematic if different methods are used across facilities, similar to the concerns Kantor et al. (2021) raised about bias in recovery controls. PMMoV concentrations were resilient to pasteurization and to freezing of the raw influent, particularly when compared to the SARS-CoV-2, which is likely due to the biology of the PMMoV as a non-enveloped tobamovirus (Alsonso et al. 1991) compared to the enveloped coronaviruses (Zhu et al. 2020). In contrast, the resilience and abundance of PMMoV in sewage is what makes it a good sewage marker and endogenous control (Kitajima et al. 2018). This is consistent with some studies that also reported differentiation in SARS-CoV-2 virus and PMMoV in influent processing (e.g. Whitney et al. 2021), while another study found the two viruses responded consistently to freezing in primary sludge (Simpson et al. 2021).

Finally, our results indicate that there is considerable difference in sensitivity among methods, which is particularly relevant as we enter a new phase of WBS that focuses on tracking low concentrations of the virus. Until now, the principal advantage of WBS has been speed, providing information a few days sooner than infectivity of individuals. However, the number of clinical testing sites and the inclination of individuals to get tested is declining as vaccination becomes widespread, reducing the reliability of individual infectivity, which has been a primary metric for public health management information. WBS has the potential to become a more reliable indicator of whether there are upticks in prevalence that results as businesses more fully reopen and use of masks and other non-pharmaceutical interventions declines. Addressing this need requires that WBS employ methods sensitive enough to detect low concentrations, rather than producing non-detects.

## Data Availability

Data is available from the authors upon request

