## Supplemental Tables for "Sources of variability in methods for processing, storing, and concentrating SARS-CoV-2 in influent from urban wastewater treatment plants"

**Supplemental Material**

Supplemental Tables

Table S1. Description of wastewater treatment facilitates utilized for each experiment. n refers to number of samples collected and processed from each WTF.

| **Experiment** | **Facility** | **Location** | **Facility ID** | **n** |
| --- | --- | --- | --- | --- |
| Sample Storage and Preservation | Hyperion Treatment Plant | Playa del Rey, CA | WTP A | 3 |
|  | Orange County Treatment Plant #1 | Fountain Valley, CA | WTP D | 3 |
|  | Point Loma Wastewater Treatment Plant | San Diego, CA | WTP E | 3 |
| Heat Inactivation of Viruses | Hyperion Treatment Plant | Playa del Rey, CA | WTP A | 3 |
|  | Joint Water Pollution Control Plant | Carson, CA | WTP B | 3 |
|  | San Jose Creek Water Reclamation Plant | Whittier, CA | WTP C | 3 |
| Virus Capture and RNA Extraction | Hyperion Treatment Plant | Playa del Rey, CA | WTP A | 6 |
|  | Joint Water Pollution Control Plant | Carson, CA | WTP B | 3 |
|  | San Jose Creek Water Reclamation Plant | Whittier, CA | WTP C | 3 |
|  | Orange County Treatment Plant #1 | Fountain Valley, CA | WTP D | 3 |
|  | Point Loma Wastewater Treatment Plant | San Diego, CA | WTP E | 3 |
| SARS-CoV-2 Gene Target Selection | Hyperion Treatment Plant | Playa del Rey, CA | WTP A | 169 |
|  | Orange County Treatment Plant #1 | Fountain Valley, CA | WTP D | 29 |
|  | Point Loma Wastewater Treatment Plant | San Diego, CA | WTP E | 98 |
| RT-qPCR to ddPCR Comparison | Hyperion Treatment Plant | Playa del Rey, CA | WTP A | 30 |

Table S2 Primers and Probes

| Assay Name | Sequence | Reference |
| --- | --- | --- |
| 2019-nCoV_N1 CDC | 2019-nCoV_N1-F CDC1 GACCCCAAAATCAGCGAAAT  2019-nCoV_N1-R CDC1 TCTGGTTACTGCCAGTTGAATCTG  2019-nCoV_N1-P CDC1 FAM-ACCCCGCATTACGTTTGGTGGACC-BHQ1 | Lu et al. 2020 |
| 2019-nCoV_N2 CDC | 2019-nCoV_N2-F TTACAAACATTGGCCGCAAA  2019-nCoV_N2- GCGCGACATTCCGAAGAA  2019-nCoV_N2-P SUN-ACAATTTGCCCCCAGCGCTTCAG-BHQ1 | Lu et al. 2020 |
| Bovine-CoV | Bovine-CoV-Forward C+TGGAAGTTGGTGGAGTT  Bovine-CoV-Reverse ATTATCGG+CCTAACATAC+ATC  Bovine-Covid-Probe /5HEX/ACCCAGAAA/ZEN/CAAACAACTTGATGTGTATAGATATGAA/3IABkFQ/ | Decaro et al. 2008 |
| PMMoV | PMMoV-Forward GAGTGGTTTGACCTTAACGTTGA  PMMoV-Reverse TTGTCGGTTGCAATGCAAGT  PMMoV-Probe-Hex (LNA) /5HEX/C+CTA+CC+GAAG+CA+AATG/3IABkFQ/QA/ | Kitajima et al. 2018, Gonzalez et al. 2020 |
